# Clinical encounter heterogeneity and methods for resolving in networked EHR data: A study from N3C and RECOVER programs

**DOI:** 10.1101/2022.10.14.22281106

**Authors:** Peter Leese, Adit Anand, Andrew Girvin, Amin Manna, Saaya Patel, Yun Jae Yoo, Rachel Wong, Melissa Haendel, Christopher G Chute, Tellen Bennett, Janos Hajagos, Emily Pfaff, Richard Moffitt

## Abstract

**OBJECTIVE:** Clinical encounter data are heterogeneous and vary greatly from institution to institution. These problems of variance affect interpretability and usability of clinical encounter data for analysis. These problems are magnified when multi-site electronic health record data are networked together. This paper presents a novel, generalizable method for resolving encounter heterogeneity for analysis by combining related atomic encounters into composite ‘macrovisits.’

**MATERIALS AND METHODS:** Encounters were composed of data from 75 partner sites harmonized to a common data model as part of the NIH Researching COVID to Enhance Recovery Initiative, a project of the National Covid Cohort Collaborative. Summary statistics were computed for overall and site-level data to assess issues and identify modifications. Two algorithms were developed to refine atomic encounters into cleaner, analyzable longitudinal clinical visits.

**RESULTS:** Atomic inpatient encounters data were found to be widely disparate between sites in terms of length-of-stay and numbers of OMOP CDM measurements per encounter. After aggregating encounters to macrovisits, length-of-stay (LOS) and measurement variance decreased. A subsequent algorithm to identify hospitalized macrovisits further reduced data variability.

**DISCUSSION:** Encounters are a complex and heterogeneous component of EHR data and native data issues are not addressed by existing methods. These types of complex and poorly studied issues contribute to the difficulty of deriving value from EHR data, and these types of foundational, large-scale explorations and developments are necessary to realize the full potential of modern real world data.

**CONCLUSION:** This paper presents method developments to manipulate and resolve EHR encounter data issues in a generalizable way as a foundation for future research and analysis.

## BACKGROUND AND SIGNIFICANCE

While the terms “encounter” and “visit” are used interchangeably to describe many different types of experiences in healthcare, these terms represent a much more specific concept within electronic health records (EHRs). In EHR data, the encounter is a transactional unit of health service delivery. Encounters can represent single, discrete health care services, such as an outpatient office visit; multiple unrelated care services, such as multiple outpatient events over a single or multiple days; or multiple related care events over short or long time periods, such as the variety of facility and professional services delivered during hospitalization. This tendency of encounters to function as the building blocks of larger and more complex care events contributes to their complexity, particularly when assessing patient care longitudinally. Unfortunately, methods to associate encounters into complete, clinically-recognizable care experiences are neither proscribed, straightforward, nor harmonized between different healthcare organizations, different EHR platforms, or even the same EHR platform implemented at different sites [[1–4]]. This data variation is the result of the significant variation in clinical service delivery that has been and continues to be widely documented[[5-7]], and also the equivalent variation in EHR implementation, utilization and clinical data documented and produced[8,9]. Unless resolved (generally on a project-by-project basis), this heterogeneity and ambiguity can undermine the encounter’s value in analysis and is a major obstacle to performing accurate, reliable analysis of EHR.

While working with encounters in the EHR is challenging within a single institution the lack of recognized, shared standards for the encounter concept causes even greater harmonization and analytical issues in multi-institution, EHR-based research. Even when participating institutions use the same common data model (CDM, such as OMOP, PCORnet, or i2b2/ACT), each of which have mechanisms for incorporating encounters,[[10–12]] none of the CDMs enforce common rules or definitions for what events constitute an encounter and how related encounters should be linked. Thus, when an institution populates a CDM with their EHR data, that institution is typically, simply translating their local definition(s) for encounters into the CDM, where these data become “harmonized” to the CDM but remain unstandardized to other encounters. As an example, one site may break inpatient encounters into a series of separate, discrete short encounters, and another may use one encounter record for the entire inpatient stay.

While many types of encounters are complex, hospitalizations (including inpatient, observation, extended recovery, and other longitudinal facility-based encounters) are most affected by encounter variation. Hospitalizations typically span a longer temporal period and tend to include a greater number of services and resources than outpatient encounters. At a minimum, hospitalizations require the combination of both facility and professional transactions to capture the full care experience. It is therefore common for hospitalizations to include many discrete encounter records to capture a wide variety of activities occurring during the hospitalization, such as imaging, pharmacy, surgery, and other services. A distinct challenge when working with encounter data for hospitalizations is cleanly identifying each discrete hospitalization from admission to discharge with all related, co-occurring services. EHR applications tend to solve this problem by having tables and methods separate from encounters to account for entire hospitalizations, such as bundled ‘accounts’ or ‘episodes’. However, these EHR-native methods are not currently represented outside of EHR platforms, and are notably missing in CDMs which frequently serve as data exchange standards for research. This means the most common research situation is that data users must attempt to identify and re-aggregate the components of hospitalizations on their own.

The challenge of working with heterogeneous EHR encounter data can be practically illustrated by the National COVID Cohort Collaborative (N3C), which networks EHR data from 75 sites and four CDMs (OMOP, PCORnet, i2b2/ACT, TriNetX) into a single repository to support community-driven, reproducible, and transparent COVID-19 analytics[13]. Encounter-level data, particularly around hospitalizations, are highly desirable for COVID-19 and post-acute sequelae of SARS-CoV-2 (PASC) research; however, early in the process of assessing N3C data quality, several obstacles became apparent in the combined, harmonized visit data from participating sites. One issue is the combination of heterogeneous local encounter definitions such as the prior described differences in recording inpatient stays. Additionally, some sites are likely mismapping a subset of their encounters to CDM visit types at the local level, such as incorrectly mapping facility-based outpatient encounters to an inpatient visit type. These issues result in encounter data that are difficult to analyze holistically, and greatly impact the ability of researchers to quantify and assess events occurring during hospitalizations. For this reason, the N3C community recognized the need to create an algorithmic method to collapse concurrent encounters for the same patient into a single analytical unit, approximating an aggregated care experience inclusive of all services.

## OBJECTIVE

Local business rules defining inpatient encounters are entrenched in their home organizations, and are often put in place for pragmatic or business optimization purposes. For this reason, it is unlikely that encounters (inpatient or otherwise) can be fully standardized from the individual organizations’ EHRs, short of the creation of national health data standards. The objectives of this work are then 1) to identify and describe the heterogeneity of harmonized CDM encounter data in the context of hospitalizations, 2) to enumerate example algorithms for re-combining transactional EHR encounters *post hoc* into logical, longitudinal care experiences with acceptable metadata characteristics, and 3) to examine the results of applying these algorithms in N3C. This process of combining atomic encounters back into longitudinal clinical experiences representative of the patient clinical experience, which we termed *macrovisit aggregation*, can be applied to an encounter dataset composed of mixed local definitions, and result in a consistently defined set of longitudinal, multi-encounter experiences, *macrovisits*, for use in further analyses.

## MATERIALS AND METHODS

### Data & Technology

This study is part of the NIH Researching COVID to Enhance Recover (RECOVER) Initiative, which seeks to understand, treat, and prevent PASC. For more information on RECOVER, visit https://recovercovid.org.

The design of the N3C OMOP data repository, the N3C data transformation pipeline, and a comprehensive characterization of the data available prior to December 2020 have been previously described[[14,15]]. In the current study, we used N3C data ingested as of 8/26/2022, which included 75 contributing sites, 15,231,849 distinct patients, and 894,629,506 encounter records. All technical work was performed in the N3C secure data enclave utilizing the Foundry technology platform created and maintained by Palantir Technologies Incorporated. All data engineering and analysis was performed using a combination of SQL, R version 3.5, and Python version 3.6.

### The Macrovisit Aggregation Algorithm

The macrovisit aggregation algorithm aims to combine individual OMOP visit records (“microvisits”) that appear to be part of the same care experience, and create a single macrovisit to represent the entirety of the care experience. Events occurring during any microvisit can then be analyzed in the context of the macrovisit rather than as individual, unrelated encounters.

In short, macrovisit aggregation merges overlapping microvisits with specific features to determine the total macrovisit’s duration; subsequently, any microvisits occurring within the timespan of the macrovisit duration are appended. Microvisits from individual sites are sourced from N3C’s visit_occurrence table. Microvisits qualified to initiate macrovisit aggregation meet the following criteria:

- have non-null start and end dates
- have a non-negative LOS, the difference between the visit’s end date and start date
- have a recorded visit_type_concept of one of the following OMOP concepts: 262 “Emergency Room and Inpatient Visit,” 8717 “Inpatient Hospital,” 9201 “Inpatient Visit,” 32037 “Intensive Care,” 581379 “Inpatient Critical Care Facility”

In addition to these inpatient-centric macrovisits, certain longitudinal, outpatient facility stays can generate macrovisits; namely, microvisits with LOS >= 2 days and type 9203 “Emergency Room Visit,” 8756 “Outpatient Hospital,” or 581385 “Observation Room.”

**Figure 1**. illustrates macrovisit aggregation with example microvisit data.

**Figure 1:**
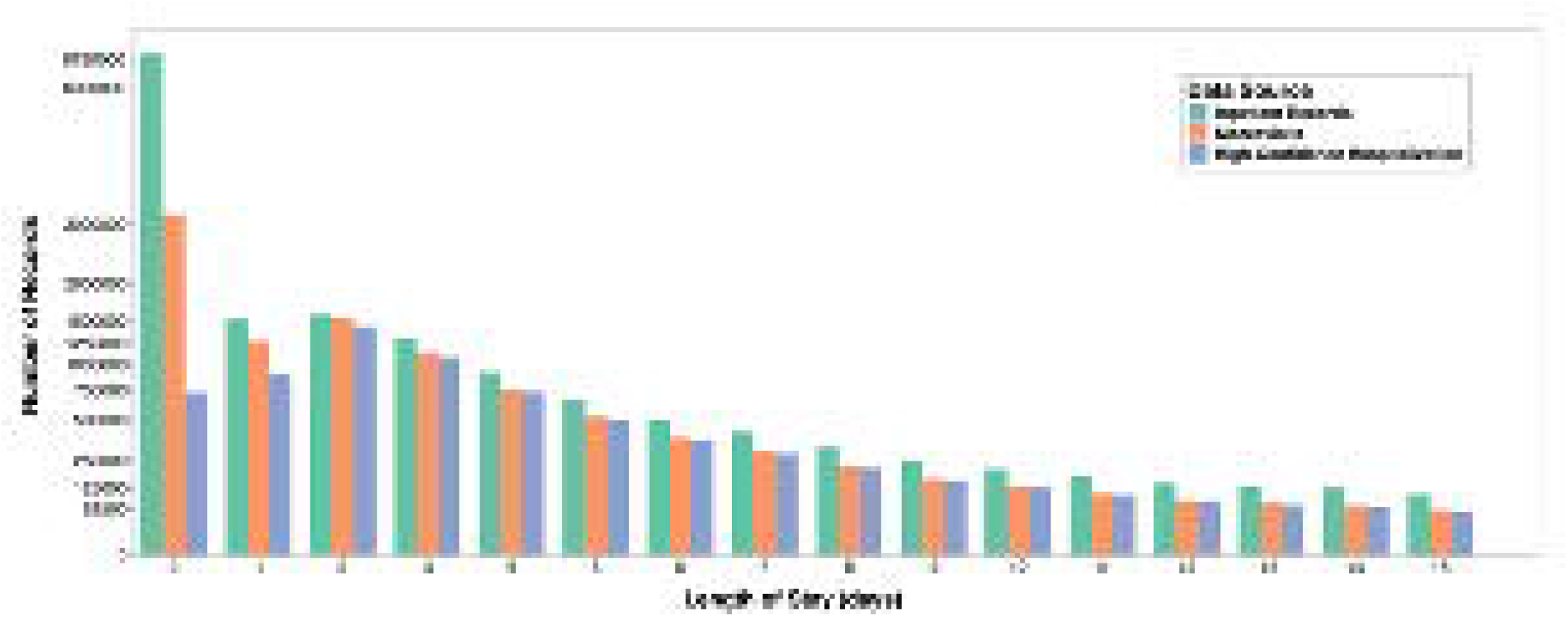
Visual representation of macrovisit aggregation. Overlapping microvisits are merged into a continuous, bundled macrovisit and the earliest visit start date and latest visit end date are assigned to the macrovisit. Microvisits that occur >=1 calendar day later with no other, overlapping microvisit generate distinct macrovisits.

### The High-Confidence Hospitalization Algorithm

Initial analysis of macrovisit aggregation output showed an unusually high frequency of 0-day macrovisits, suggesting a substantially larger number of extremely short hospitalizations, and indicating a hospitalization length-of-stay deviating from the expected distribution[[16–18]]. The typical expected distribution of hospital length-of-stay for an acute care hospital is approximately Poisson or negative binomial distributed with a central tendency around 2 to 4 days. This indicated that a macrovisit was not equivalent to a hospitalization in all cases. There are several possible explanations for this finding, including mislabelling of outpatient visits by sites as inpatient type, mislabeling a brief inpatient service (such as critical care) as an entire hospitalization without sending additional data, or inaccurately recording the visit start and/or end date. It became clear that additional filtering was required to further classify macrovisits into the categories of “high-confidence hospitalization” and “non-hospitalization macrovisits.” Due to the heterogeneity of visit data submitted between the many N3C sites, an ensemble of approaches was constructed to attempt to perform this classification, independent of either LOS or site-submitted visit_concept_types. Criteria included:

- Presence of diagnosis-related group (DRG) codes for any component microvisit OR
- Presence of Centers for Medicare & Medicaid Services (CMS)-indicated inpatient-only Current Procedural Terminology (CPT) codes [[19]] on any component microvisit
OR
- Presence of either an inpatient or critical care (ICU) evaluation & management Healthcare Common Procedure Coding System (HCPCS) code on any component microvisit
OR
- Presence of either an inpatient or ICU SNOMED CT concept on any microvisit procedure
OR
- A minimum of 50 total *resources* recorded for at least 1 component microvisit (where total resources consists of the total count of all diagnoses, procedures, medications, measurements, and observations at the microvisit level)

The resources attributed to each macrovisit (hereafter ‘resource density’) LOS are illustrated in **Figure 2**. Examination of all these indicators simultaneously facilitated identifying macrovisits with either inpatient hospital care delivery or with a resource pattern consistent with a likely longitudinal hospital encounter.

**Figure 2:**
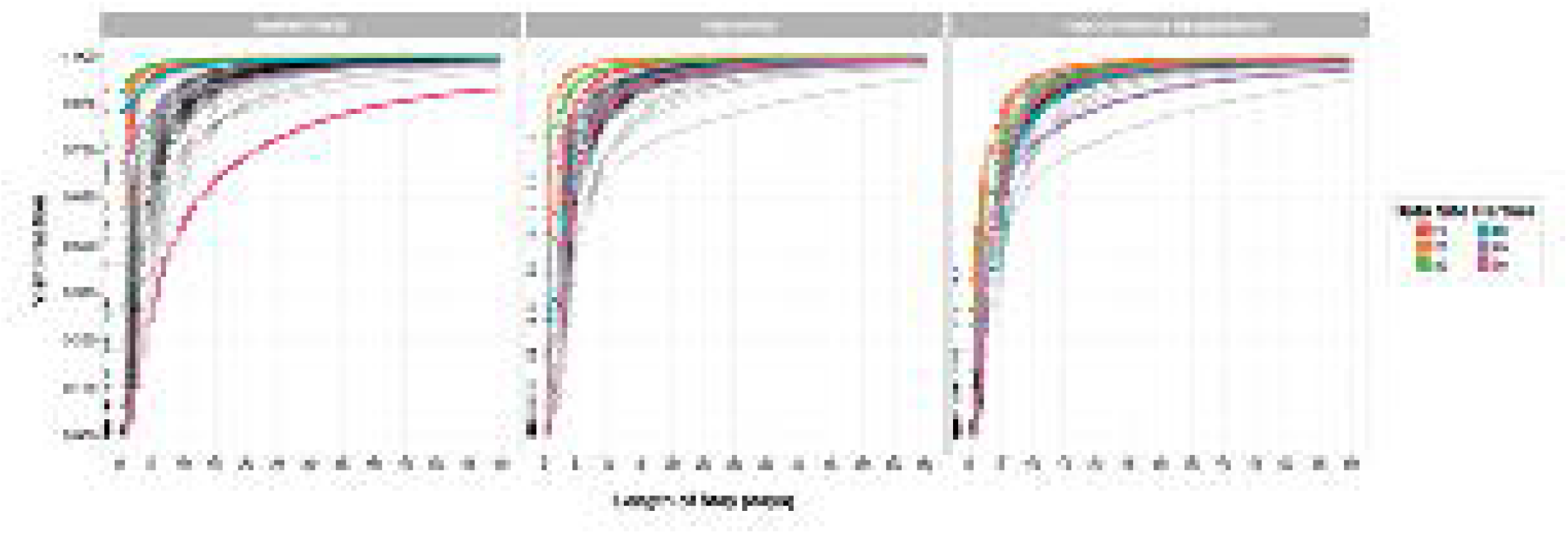
Resource density by length of stay. Visual representation of the variation in maximum resource density across macrovisits as LOS changes. Each color corresponds to a maximum resource density bin. The macrovisits with greater than 50 maximum resources roughly follow the expected inpatient LOS distribution, and a majority of 0-day LOS macrovisits have no more than 25 maximum resources.

### Assessing Site-to-Site Encounter Heterogeneity

To assess the performance of the macrovisit aggregation algorithm, the characteristics of microvisits labeled as inpatient (visit types 262 “Emergency Room and Inpatient Visit,” 8717 “Inpatient Hospital,” 9201 “Inpatient Visit,” 32037 “Intensive Care,” or 581379 “Inpatient Critical Care Facility”) from the visit_occurrence table were compared to the generated macrovisits across all N3C sites. Measurement (defined as data from the OMOP measurements table, consisting of labs, vitals, and other structured quantitative clinical assessments) frequency is included to illustrate the data density compiled into the macrovisits from the component microvisits and to identify data quality issues, such as long macrovisits with a small number of associated measurements. The goal was to ensure that the algorithm behaved similarly across sites and decreased heterogeneity. The workflow for assessing performance is detailed in **Table 1**.

**Table 1.**
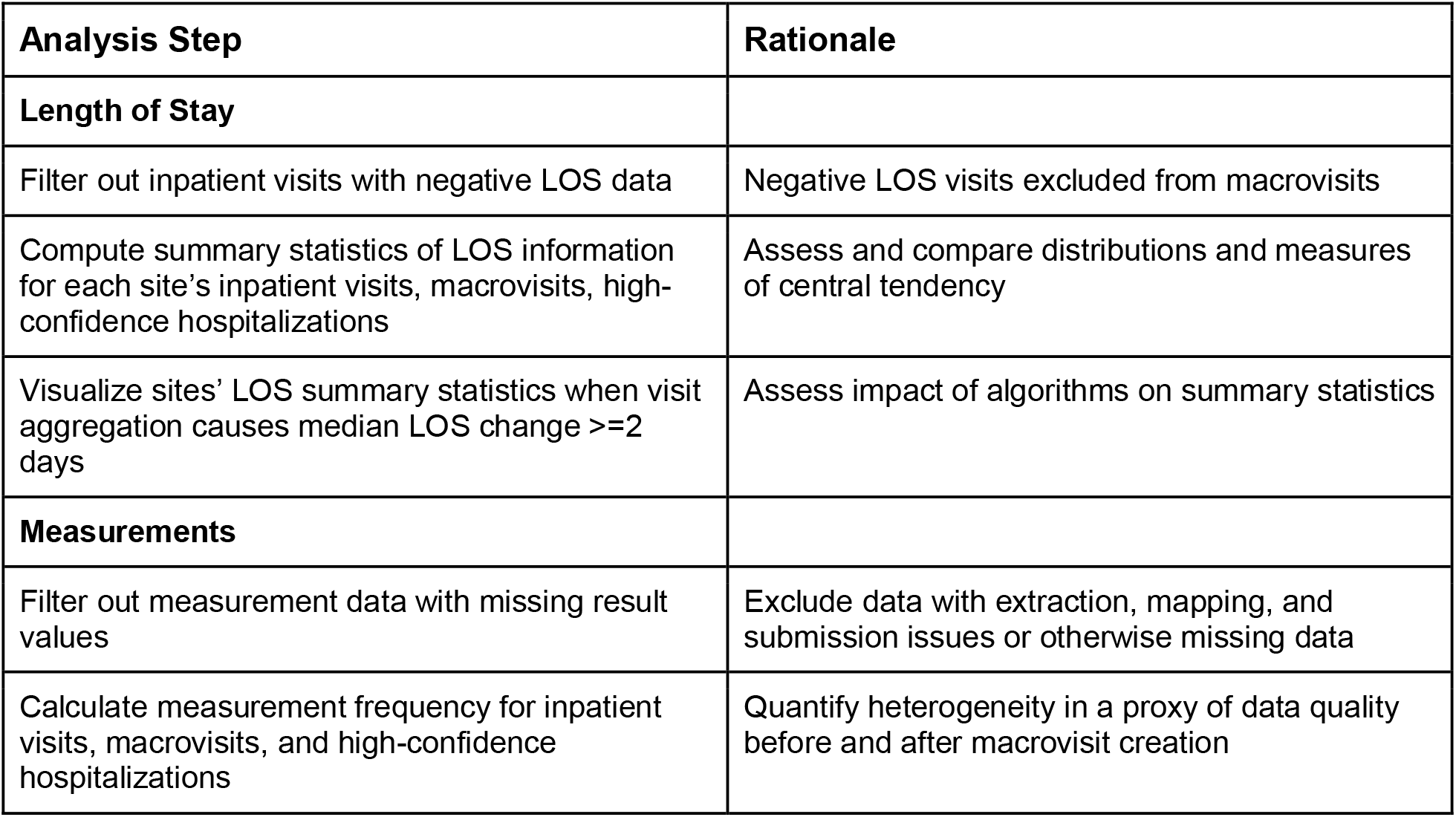
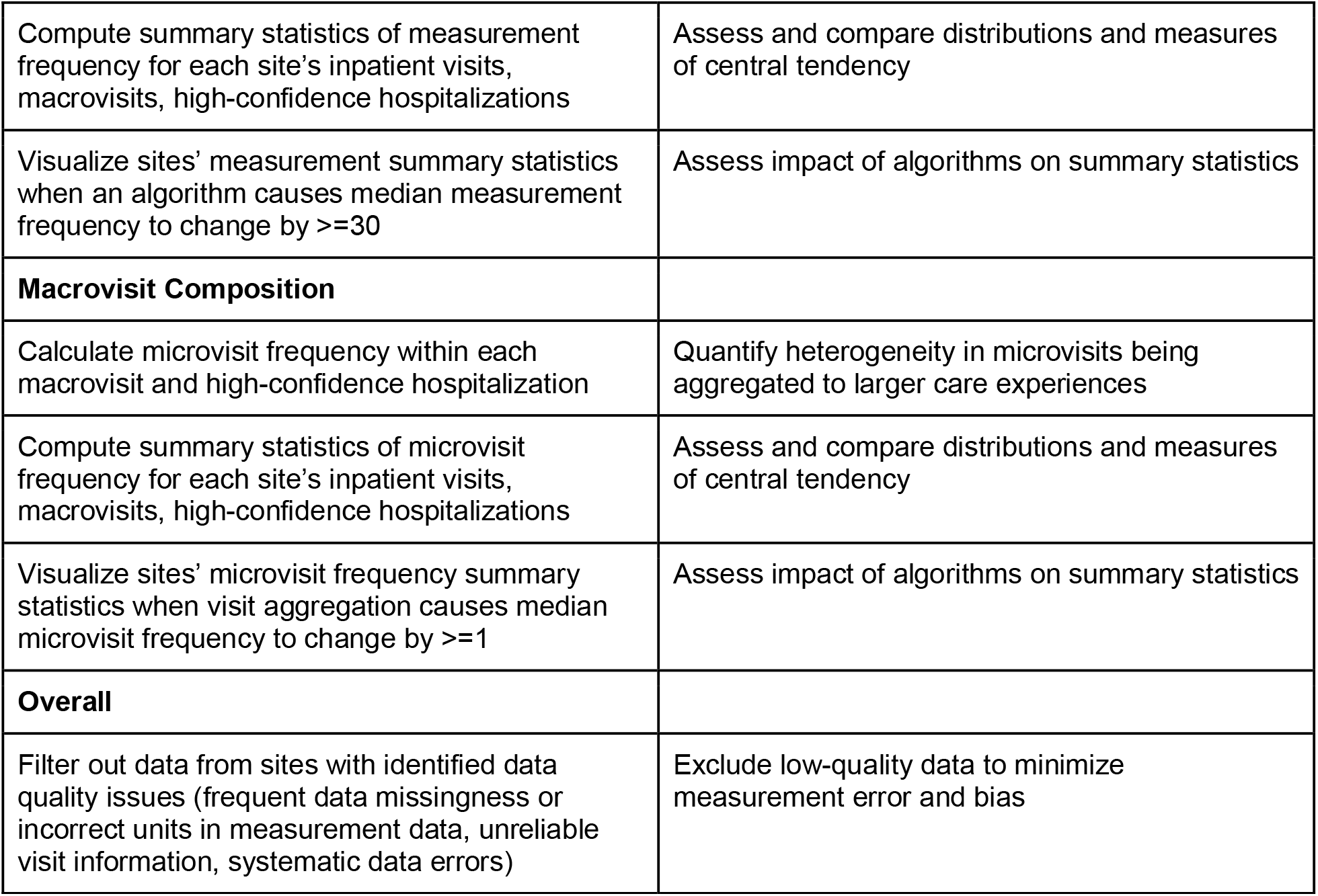
Steps taken to asses site-level encounter heterogeneity

| **Analysis Step** | **Rationale** |
| --- | --- |
| **Length of Stay** |  |
| Filter out inpatient visits with negative LOS data | Negative LOS visits excluded from macrovisits |
| Compute summary statistics of LOS information for each site’s inpatient visits, macrovisits, high-confidence hospitalizations | Assess and compare distributions and measures of central tendency |
| Visualize sites’ LOS summary statistics when visit aggregation causes median LOS change $\geq 2$ days | Assess impact of algorithms on summary statistics |
| **Measurements** |  |
| Filter out measurement data with missing result values | Exclude data with extraction, mapping, and submission issues or otherwise missing data |
| Calculate measurement frequency for inpatient visits, macrovisits, and high-confidence hospitalizations | Quantify heterogeneity in a proxy of data quality before and after macrovisit creation |
| Compute summary statistics of measurement frequency for each site's inpatient visits, macrovisits, high-confidence hospitalizations | Assess and compare distributions and measures of central tendency |
| Visualize sites' measurement summary statistics when an algorithm causes median measurement frequency to change by $\geq 30$ | Assess impact of algorithms on summary statistics |
| **Macrovisit Composition** |  |
| Calculate microvisit frequency within each macrovisit and high-confidence hospitalization | Quantify heterogeneity in microvisits being aggregated to larger care experiences |
| Compute summary statistics of microvisit frequency for each site's inpatient visits, macrovisits, high-confidence hospitalizations | Assess and compare distributions and measures of central tendency |
| Visualize sites' microvisit frequency summary statistics when visit aggregation causes median microvisit frequency to change by $\geq 1$ | Assess impact of algorithms on summary statistics |
| **Overall** |  |
| Filter out data from sites with identified data quality issues (frequent data missingness or incorrect units in measurement data, unreliable visit information, systematic data errors) | Exclude low-quality data to minimize measurement error and bias |

## RESULTS

### Macrovisit Composition Heterogeneity

To illustrate variance in site-level encounter definitions, **Figure 3** shows a sampling of macrovisit composition across N3C sites. Each facet represents a single, randomly selected macrovisit while the colored bars indicate the variety and duration of component microvisits making up the macrovisit. It is worth noting that the microvisits labeled as ‘office visit’ or ‘outpatient visit’ might logically represent the professional component of facility care delivery instead of true, discrete ambulatory visits; however, that cannot be determined conclusively. Similarly, it is not possible to determine what true visit types might be represented by the abundance of microvisits labeled as “no matching concept” in the OMOP vocabulary. Despite these unknowns, it is clear that, as expected, macrovisits are diverse and represent a wide variety of encounter types and durations over longitudinal care.

**Figure 3:**
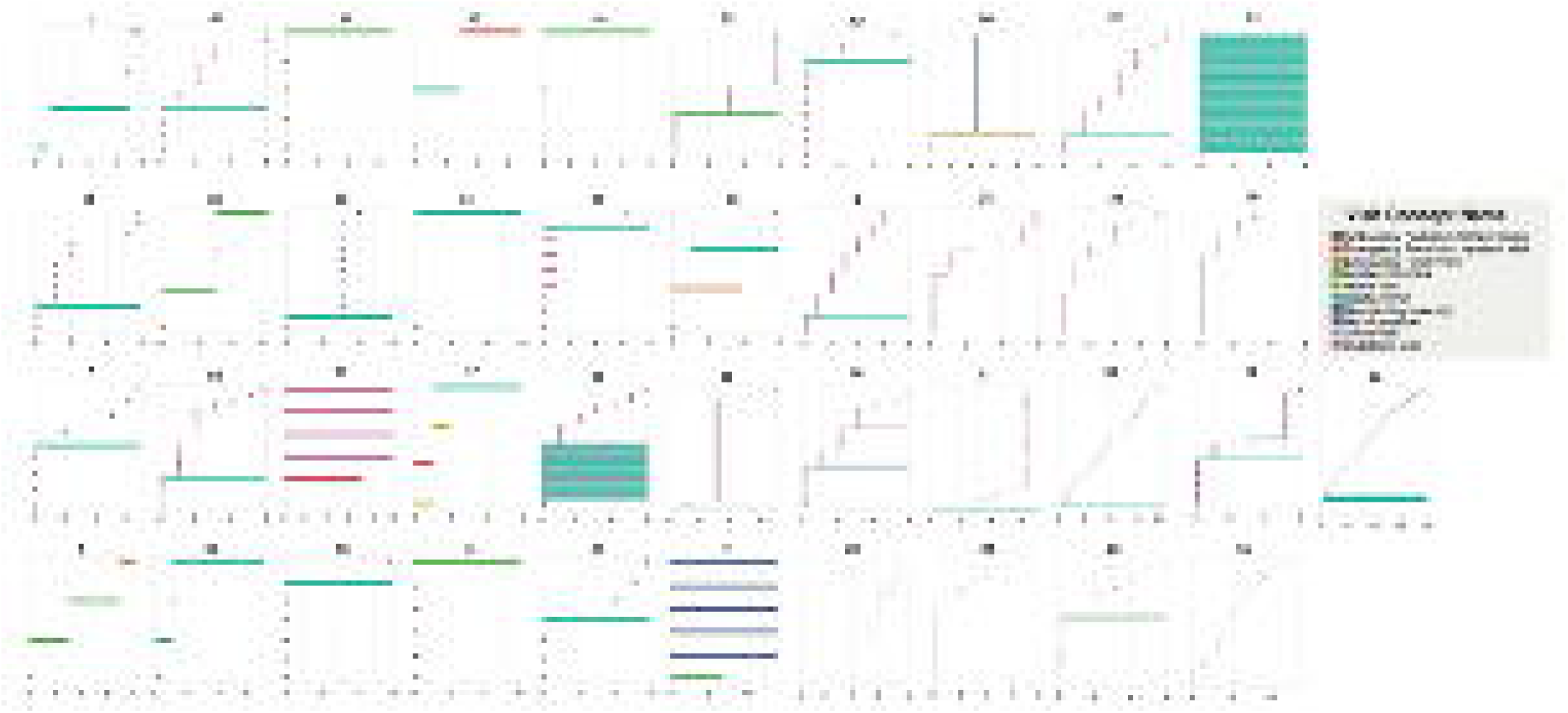
Microvisit heterogeneity within macrovisits. Visual representation of varied composition of a randomly-selected macrovisit for each site. Small colored markers indicate individual microvisits included in the macrovisit. For example, site 37 has a macrovisit consisting of one longitudinal inpatient visit with a variety of 0-day visits spread throughout the stay. Site 8 has a macrovisit consisting of 2 overlapping inpatient stays, again with a variety of 0-day visits over the entire macrovisit.

### Assessing Algorithm Impact

In addition to the composition of macrovisits, the impact of algorithms was also assessed by examining LOS, measurement frequency, and microvisit frequency within macrovisits. As illustrated previously when assessing composition in Figure 3, microvisit frequency is an important exploratory metric to understand the makeup of macrovisits and the underlying visit data heterogeneity. LOS is also a vital metric to identify the algorithm’s success in aggregating microvisits into plausible longitudinal stays that are usable for other analyses. The results of these assessments are shown in **Table 2**.

**Table 2.** Summary statistics of measured features for inpatient microvisits and both algorithms.

|  | Feature | p1 | Q1 | median | mean (sd) | Q3 | p99 |
| --- | --- | --- | --- | --- | --- | --- | --- |
| **Inpatient Visit**<br>n = 16,421,633 | Length of stay | 0 | 0 | 1 | 5.4 (19.3) | 5 | 61 |
| **Macrovisit**<br>n = 10,577,329 |  | 0 | 0 | 2 | 4.5 (12.4) | 5 | 44 |
| **High-Confidence Hospitalization**<br>n = 7,434,312 |  | 0 | 2 | 3 | 5.9 (13.8) | 6 | 49 |
| **Inpatient Visit** | Measurements per longitudinal stay | 1 | 14 | 58 | 192.4 (750.6) | 166 | 2,150 |
| **Macrovisit** |  | 1 | 27 | 82 | 244.3 (1288.7) | 205 | 2,646 |
| **High-Confidence Hospitalization** |  | 3 | 44 | 103 | 279.1 (1,378.5) | 237 | 2,904 |
| **Macrovisit** | Microvisits per macrovisit | 1 | 1 | 2 | 5.6 (14.9) | 6 | 48 |
| High-Confidence Hospitalization |  | 1 | 1 | 3 | 7.0 (17.5) | 7 | 58 |

### Length-of-Stay Impact

From Table 2 and supplemental Figure 1 it is apparent that both inpatient microvisits and macrovisits have a large proportion of zero-day care experiences, which would be unusual in true hospitalizations. In the N3C inpatient visits data, over 40% of all visits are reported as zero-day inpatient visits, which is extremely dissimilar from clinical practice. The characteristics of LOS are improved somewhat by the macrovisit algorithm as the median moves from 1 to 2 days; however, zero-day hospitalizations are still over-represented. Applying the high-confidence hospitalization algorithm moves the LOS distribution further to the right, bringing the median to 3 days. These effects are illustrated in **Figure 4** for a subset of sites, showing the increase in LOS from visits to macrovisits to high-confidence hospitalizations. Additional LOS data is shown in supplemental Figures 1 and 2.

**Figure 4:**
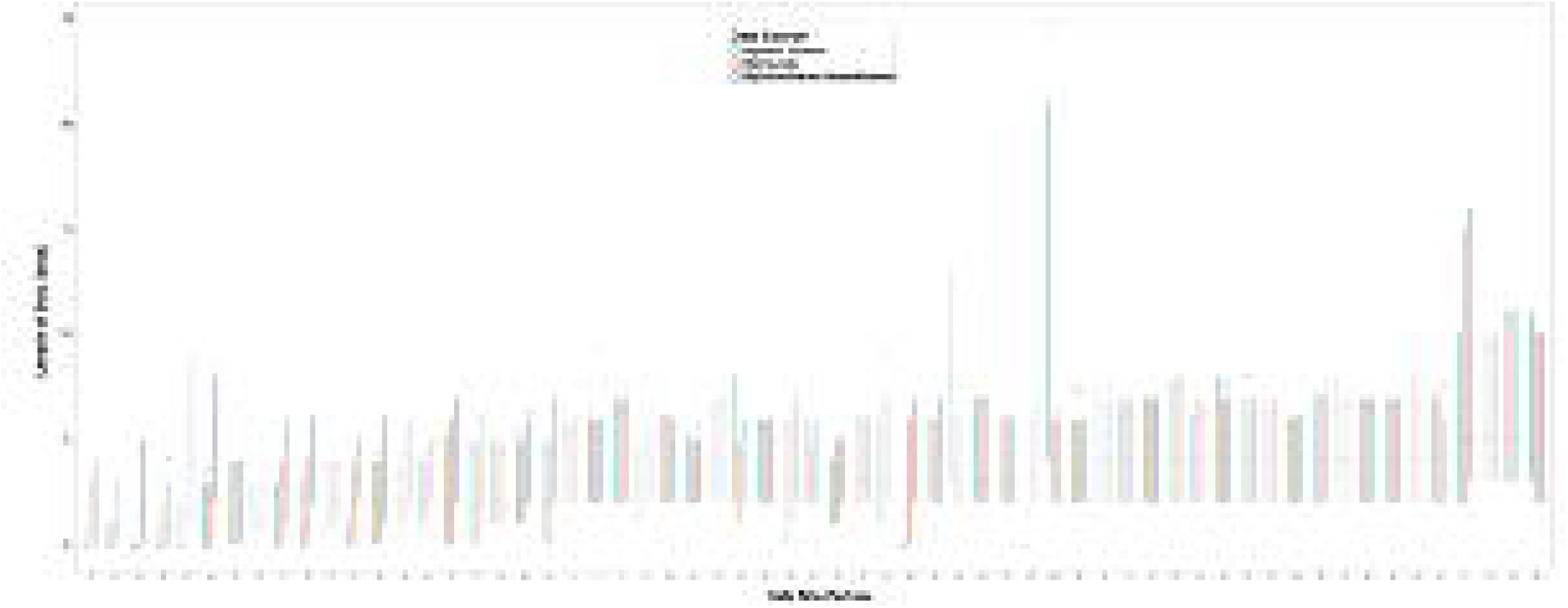
Length-of-stay. LOS distributions for inpatient visits, macrovisits, and high-confidence hospitalizations for the subset of sites with most variance between raw data and algorithm results (median=circle, mean=triangle, IQR=line).

### Microvisit Frequency Impact

The fundamental composition of macrovisits was explored by examining the frequency of microvisits within macrovisits. The mean microvisits per macrovisit were 5.6 and 7.0 for base macrovisits and high-confidence hospitalizations, respectively. Similarly, the median shifted from 2 to 3 from macrovisits to hospitalizations, collectively indicating a small increase in microvisit density from the macrovisit algorithm to the high-confidence hospitalization algorithm. Both sets of data retain a sizable amount of heterogeneity in microvisit composition with a standard deviation of 14.9 and 17.5, respectively for macrovisits and high-confidence hospitalizations. These data are illustrated for a subset of sites in **Figure 5**, showing the interquartile range, mean, and median of microvisit frequency for both macrovisits and high-confidence hospitalizations. The overall tendency of the algorithms to create increasingly microvisit-dense macrovisits is apparent from this figure.

**Figure 5:**
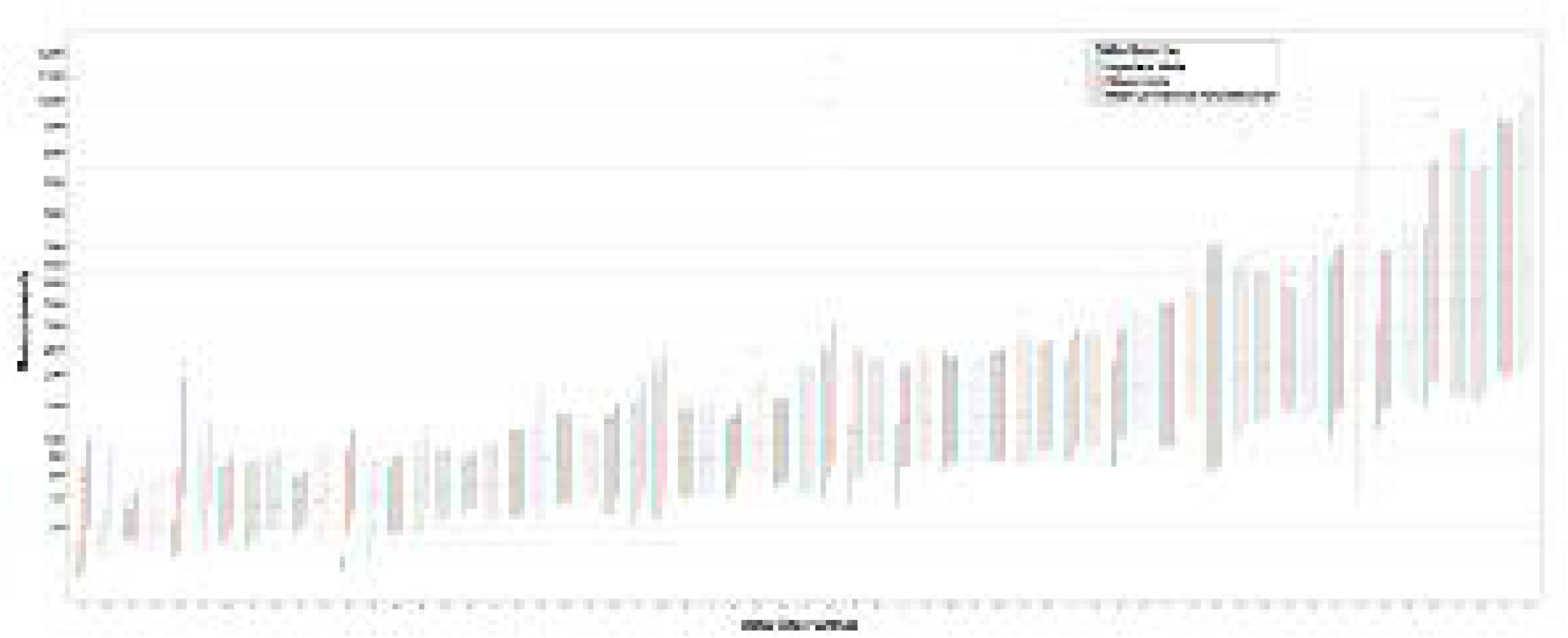
Microvisit density. Distribution of the number of component microvisits within macrovisits and high-confidence hospitalizations for the subset of sites with most variance between macrovisit algorithm and high-confidence hospitalization algorithm results (median=circle, mean=triangle, IQR=line).

### Measurement Frequency

OMOP measurements data was explored for macrovisits as a proxy of overall clinical data contained within each macrovisit. As the macrovisit and high-confidence hospitalization concepts should represent longitudinal care experiences rich with clinical data compared to single visits, measurement frequency per macrovisit is a valuable quality indicator of the function of the algorithms. This general trend is apparent as the mean measurements per inpatient visits is 192.3 compared to 244.3 and 279.1 for broad macrovisits and high-confidence hospitalizations, respectively. Similarly, the medians for these groups increase from 58 to 82 to 103, illustrating the rightward shift of the distributions and overall increasing density of measurements data. Measurement frequency data are shown in **Figure 6**, showing the site-level IQR, mean, and median for sites with the most variation from raw visits to macrovisits and hospitalizations.

**Figure 6:**
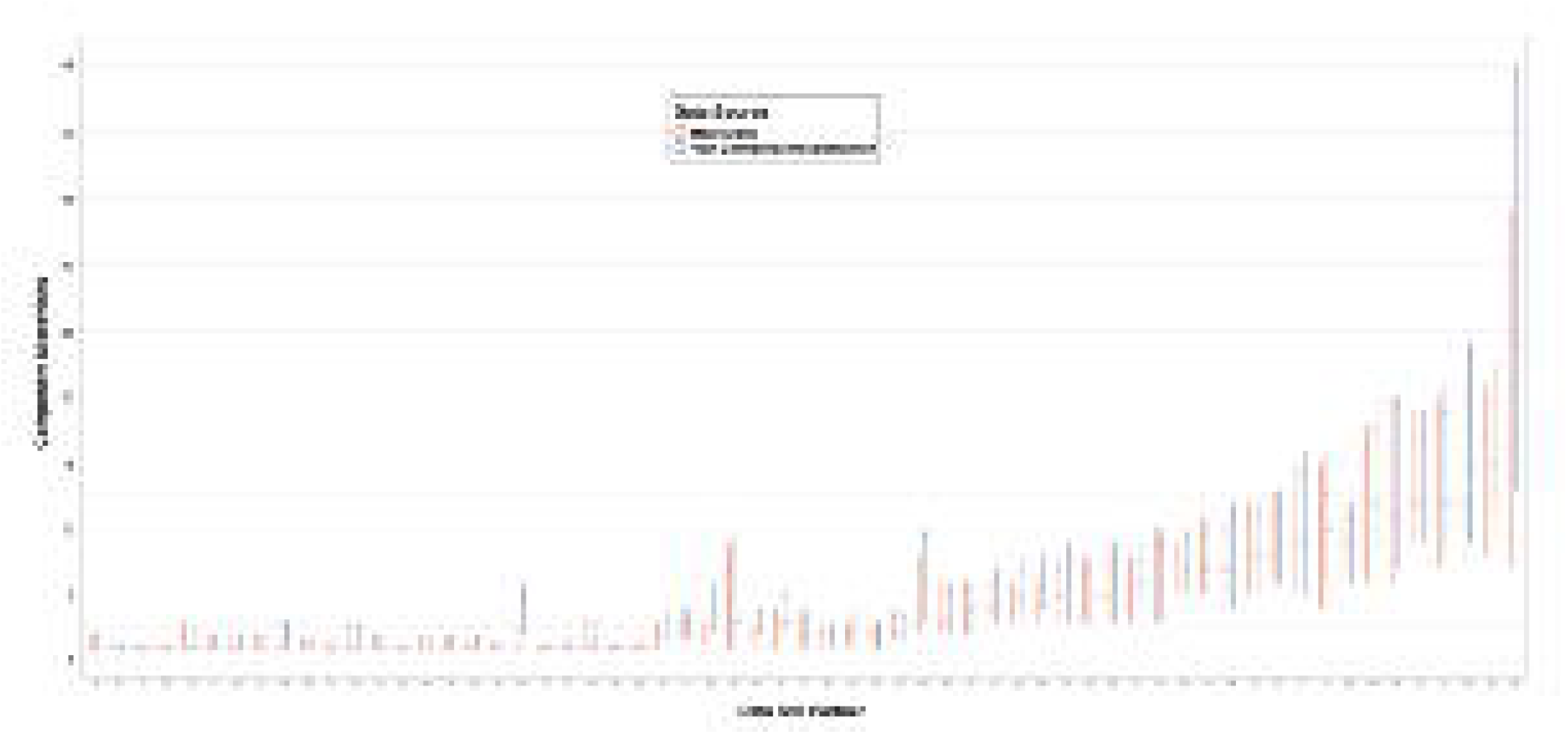
Measurement Density. Distribution of the number of measurements within inpatient visits, macrovisits, and high-confidence hospitalizations for the subset of sites with most variance between raw data and algorithm results (median=circle, mean=triangle, IQR=line).

### Clinical Application of Macrovisits to ECMO

In addition to assessing the overall, dataset-wide characteristics previously examined, macrovisit utility was also practically assessed by examining the clinical application to an extra corporeal membrane oxygenation (ECMO) cohort. ECMO is a highly invasive procedure to oxygenate the blood outside of the body, bypassing the lungs and heart and is an extremely resource intensive procedure requiring intensive care-level hospitalization. Macrovisits were assessed in ECMO cases by examining the hospital LOS and numbers of partial thrombin time(Ptt) labs performed during ECMO (Ptt is a coagulation lab required regularly when on ECMO) in **Table 3**. Examining microvisits with ECMO showed impossibly short LOS and small numbers of Ptt labs performed with medians of 0 for both. Bundling ECMO microvisits into macrovisits changes both the LOS and Ptt administration medians to 33 days and 25 Ptt labs per hospitalization with ECMO, respectively. Previous systematic review has shown mean hospital LOS for hospitalizations including ECMO to span from 12 days to 50 days.[20]

**Table 3.** Distributions of LOS and Ptt labs in ECMO visits and macrovisits. Comparing the distribution of hospital LOS and coagulation lab tests for microvisits including ECMO versus macrovisits including ECMO.

|  | 1% | 25% | 50% | 75% | 99% |
| --- | --- | --- | --- | --- | --- |
| Visit LOS<br>n=39,809 | 0 | 0 | 0 | 1 | 148 |
| Macrovisit LOS<br>n=9,830 | 0 | 16 | 33 | 59 | 282 |
| Visit Ptt<br>n=39,809 | 0 | 0 | 0 | 0 | 188 |
| Macrovisit Ptt<br>n=9,831 | 0 | 9 | 25 | 60 | 327 |

## DISCUSSION

The present study provides an illustration of the service delivery heterogeneity present in EHR data across 75 N3C partner sites in the US, and demonstrates that this heterogeneity is not resolved through the application of CDMs and data harmonization efforts alone. We have demonstrated successful use of an algorithm in addition to CDM harmonization to aggregate and classify EHR visits generated from varied, site-specific operational rules and data extraction approaches into comprehensive macrovisits more reflective of actual clinical experience. Additionally we have demonstrated that, depending on the desired outcomes, multiple successive algorithms may be necessary to parse aggregated data into data suitable for analysis.

While we refer to the macrovisit as a new concept, it is necessary to differentiate it from pre-existing clinical service aggregation methods such as bundles and care episodes. Care bundles typically refer to the linking of care services occurring over various time periods for the same underlying medical situation for the purpose of paying a limited number of bundled payments as opposed to transactional fee-for-service or DRG-based payments. Care episodes may be short and discrete, such as an episode for the care following minor trauma, or long such as the long-term range of services for chronic conditions with exacerbations, such as sickle cell anemia. In contrast to bundles and episodes, macrovisits are intended for the much more focused purpose of linking encounters together to fully represent the services experienced during a discrete hospitalization, very similarly to the intrinsic linking of encounters inside many EHR systems for actions such as facility billing. Additionally, the strategy employed in the currently described macrovisit aggregation algorithm is similar to existing strategies that have both been published and anecdotally used in other datasets[[21,22]].

While the algorithms discussed are rule-based and lack the apparent dynamism of a machine-learning based approach, they are foundational steps in formally identifying and quantifying EHR data heterogeneity within and between sites and creating generalizable solutions to resolve these issues. There is conjecture around the necessity of this type of work due to the perception that these issues will be resolved through the inherent ingestion and harmonization pipelines of CDMs or through new data interfacing paradigms, such as Health Level Seven’s Fast Healthcare Interoperability Resources (FHIR) [[23]]. FHIR accounts for the possibility of aggregating encounters with the *partOf* element in the Encounter resource. However, because *partOf* is not a required field in FHIR, it remains to be seen what proportion of FHIR-ready sites will choose to use this element and how much variation will be seen in its use. Similarly, the experience of working across N3C, the largest harmonized CDM repository in the US, has demonstrated that the CDM harmonization mechanisms currently in place are not sufficient to harmonize encounter data.

Ultimately, assessing encounter heterogeneity and methods to aggregate encounters into larger hospitalizations is important because utilizing raw visit data misleads many analyses. Notably, using raw inpatient visits to identify hospitalizations, and therefore more severe encounters, in N3C data led to an undercounting of severe cases of COVID-19. While it may be tempting to attempt to solve this issue at the source (the EHR itself), it may be more advantageous to combine visits into macrovisits *post hoc* instead, which allows for more definitional flexibility for projects and research questions with different needs. Using a *post hoc* method, the “raw” transactional visits are always available in the source data instead of destroyed in a transformation that may be upstream and opaque to the end user. This also leaves room for multiple shared macrovisit-like algorithms to serve different use cases, which, for example, may wish to preserve differences between inpatient stays and extended holds in the emergency department. It would also be valuable and worthwhile to consider CDM schema extensions to facilitate the loading of hospitalization and hospital facility data and groupings that already occur in EHR platforms, such as the “account” concept in the Epic platform. While these concepts are unlikely complete solutions to the visit issues described and would likely have their own heterogeneity both within and between sites, they offer a significantly more evolved and refined mechanism for dealing with hospitalizations from the EHR.

To put encounter and other EHR data issues into perspective, we must step back and consider the policy landscape that heralded in our current national landscape of electronic health records. A major goal of the HITECH Act, as a component of American Recovery and Reinvestment Act (ARRA), was to incentivize the adoption of EHRs at a national scale in the US - an effort which, by almost any measure, has been successful [cite]. While this evolution from paper to digital health records has arguably had some intended effect of facilitating more interoperability, data sharing, and care delivery innovations, it has also had the unintended consequence of making transparent the enormous variations in clinical care, care documentation, and EHR implementation in the US healthcare system [[24]]. From an informatics perspective these issues are manifest in the tremendous heterogeneity present in EHR data, both intra- and intersite. In the short-term following HITECH, programs such as Meaningful Use [[25]] attempted to create a standard functionality floor for EHRs by requiring such data as vitals to be able to be input and retrieved, or computerized physician order entry to be performed, but the focus of these programs was to incentivize the adoption of functional, reliable EHR platforms, not to create EHR data standards. Similarly, the private market of EHR vendors has facilitated this by allowing and supporting local customization EHR implementation and maintenance.

These issues of variation and heterogeneity have always been present and problematic locally, but have become more obtrusive as EHRs have become more prominent and multi-site clinical research networks have developed. In the N3C data enclave, the largest centralized repository of multi-site, harmonized EHR data produced to-date in the US, the full scale of these national issues is manifest. At a high level, this begs the question of how to improve EHR data and CDM ingestion and standardization to make EHR data more usable in future work. There are incremental modifications that could be made within individual sites, EHR platforms, or CDMs that begin to ameliorate these varied issues. One such suggestion would be for CDMs to require both atomic encounters from EHR data and also the EHR-native hospitalizations with keys between the two. The more systemic solution, though, would be for healthcare, like many sectors before, to adopt widespread data standards either through regulation, as in finance, or through industry-sponsored, multilateral working groups, as many standards in technology use. Until that point, an excess of time will continue to be spent on figuring out *how* to get value from healthcare data instead of getting value from healthcare data.

## Data Availability

All data used in performing the work reported in this manuscript is available trough the National Institute of Health's National Covid Cohort Collaborative (N3C) enclave.

https://covid.cd2h.org/enclave

## Limitations

In retrospect, the heterogeneity and resulting issues of visits data in N3C could possibly have been mediated by a more formal use of the OMOP visit_detail table for atomic visits data and the visit_occurrence table for modified or aggregated visits data. At the time of initial development, all OMOP-native data sites only populated the visit_occurrence table with transactional visits data, and N3C followed this data loading paradigm. While this differs from the theoretically intended use of the OMOP tables, it also represents the real world adoption and utilization of common data models in multi-site networked research. It is unrealistic to rely inherently and solely on informatics tools like common data models and ontology-mapping to resolve the tribalism and heterogeneity inherent in American healthcare and the resulting data. The current study highlights these issues and the ongoing need to have both post-hoc tools for resolving data issues and significant expertise in understanding healthcare data to utilize EHR data.

As has been well documented, missingness is a common issue with all EHR data, N3C included. When data is missing or null it is not possible to make assumptions about the data or the intent of the data provider[[26–29]]. Similarly, some data, while present, appears to be illogical or mis-mapped, which is equally difficult to interpret and use. Due to the nature of the N3C data ingestion and de-identification policies, it is not possible to validate our algorithms’ assumptions using chart review. Thus, a logical next step for this work is to perform validation by running the macrovisit algorithms on local site data--preferably a selection of sites with different local definitions for encounters.

## FUNDING

This research was funded by the National Institutes of Health (NIH) Agreement OTA OT2HL161847 as part of the Researching COVID to Enhance Recovery (RECOVER) research program, as well as CD2H - The National COVID Cohort Collaborative (N3C) U24TR002306.

## COMPETING INTERESTS

None to declare.

## AUTHOR CONTRIBUTIONS

Authorship was determined using ICMJE recommendations. The content is solely the responsibility of the authors and does not necessarily represent the official views of the National Institutes of Health, N3C, or RECOVER.

Manuscript drafting: PJL, AA, AG, SP, YJY, RW, ERP, RM

Data analysis: PJL, AA, AG, AM, SP, YJY, ERP, RM

Program leadership: MH, CGC, TB, JH, ERP, RM

Final manuscript approval: PJL, AA, AG, SP, YJY, MH, TB, ERP, RM

## DATA AVAILABILITY

The N3C Data Enclave is managed under the authority of the NIH; information can be found at ncats.nih.gov/n3c/resources. Enclave data is protected, and can be accessed for COVID-related research with an approved (1) IRB protocol and (2) Data Use Request (DUR). A detailed accounting of data protections and access tiers is found in [1]. Enclave and data access instructions can be found at https://covid.cd2h.org/for-researchers; all code used to produce the analyses in this manuscript is available within the N3C Enclave to users with valid login credentials to support reproducibility.

## ACKNOWLEDGEMENT

We would like to thank the National Community Engagement Group (NCEG), all patient, caregiver and community Representatives, and all the participants enrolled in the RECOVER Initiative.

The analyses described in this publication were conducted with data or tools accessed through the NCATS N3C Data Enclave covid.cd2h.org/enclave and supported by CD2H - The National COVID Cohort Collaborative (N3C) IDeA CTR Collaboration 3U24TR002306-04S2 NCATS U24 TR002306. This research was possible because of the patients whose information is included within the data from participating organizations (covid.cd2h.org/dtas) and the organizations and scientists (covid.cd2h.org/duas) who have contributed to the on-going development of this community resource (cite this https://doi.org/10.1093/jamia/ocaa196).

## On behalf of the N3C and RECOVER consortia

The N3C data transfer to NCATS is performed under a Johns Hopkins University Reliance Protocol # IRB00249128 or individual site agreements with NIH. The Data Use Request ID is DUR-94BBC49. The N3C Data Enclave is managed under the authority of the NIH; information can be found at https://ncats.nih.gov/n3c/resources.

We gratefully acknowledge the following core contributors to N3C:

Adam B. Wilcox, Adam M. Lee, Alexis Graves, Alfred (Jerrod) Anzalone, Amin Manna, Amit Saha, Amy Olex, Andrea Zhou, Andrew E. Williams, Andrew Southerland, Andrew T. Girvin, Anita Walden, Anjali A. Sharathkumar, Benjamin Amor, Benjamin Bates, Brian Hendricks, Brijesh Patel, Caleb Alexander, Carolyn Bramante, Cavin Ward-Caviness, Charisse Madlock-Brown, Christine Suver, Christopher Chute, Christopher Dillon, Chunlei Wu, Clare Schmitt, Cliff Takemoto, Dan Housman, Davera Gabriel, David A. Eichmann, Diego Mazzotti, Don Brown, Eilis Boudreau, Elaine Hill, Elizabeth Zampino, Emily Carlson Marti, Emily R. Pfaff, Evan French, Farrukh M Koraishy, Federico Mariona, Fred Prior, George Sokos, Greg Martin, Harold Lehmann, Heidi Spratt, Hemalkumar Mehta, Hongfang Liu, Hythem Sidky, J.W. Awori Hayanga, Jami Pincavitch, Jaylyn Clark, Jeremy Richard Harper, Jessica Islam, Jin Ge, Joel Gagnier, Joel H. Saltz, Joel Saltz, Johanna Loomba, John Buse, Jomol Mathew, Joni L. Rutter, Julie A. McMurry, Justin Guinney, Justin Starren, Karen Crowley, Katie Rebecca Bradwell, Kellie M. Walters, Ken Wilkins, Kenneth R. Gersing, Kenrick Dwain Cato, Kimberly Murray, Kristin Kostka, Lavance Northington, Lee Allan Pyles, Leonie Misquitta, Lesley Cottrell, Lili Portilla, Mariam Deacy, Mark M. Bissell, Marshall Clark, Mary Emmett, Mary Morrison Saltz, Matvey B. Palchuk, Melissa A. Haendel, Meredith Adams, Meredith Temple-O’Connor, Michael G. Kurilla, Michele Morris, Nabeel Qureshi, Nasia Safdar, Nicole Garbarini, Noha Sharafeldin, Ofer Sadan, Patricia A. Francis, Penny Wung Burgoon, Peter Robinson, Philip R.O. Payne, Rafael Fuentes, Randeep Jawa, Rebecca Erwin-Cohen, Rena Patel, Richard A. Moffitt, Richard L. Zhu, Rishi Kamaleswaran, Robert Hurley, Robert T. Miller, Saiju Pyarajan, Sam G. Michael, Samuel Bozzette, Sandeep Mallipattu, Satyanarayana Vedula, Scott Chapman, Shawn T. O’Neil, Soko Setoguchi, Stephanie S. Hong, Steve Johnson, Tellen D. Bennett, Tiffany Callahan, Umit Topaloglu, Usman Sheikh, Valery Gordon, Vignesh Subbian, Warren A. Kibbe, Wenndy Hernandez, Will Beasley, Will Cooper, William Hillegass, Xiaohan Tanner Zhang. Details of contributions available at covid.cd2h.org/core-contributors

The following institutions whose data is released or pending:

Available: Advocate Health Care Network — UL1TR002389: The Institute for Translational Medicine (ITM) • Boston University Medical Campus — UL1TR001430: Boston University Clinical and Translational Science Institute • Brown University — U54GM115677: Advance Clinical Translational Research (Advance-CTR) • Carilion Clinic — UL1TR003015: iTHRIV Integrated Translational health Research Institute of Virginia • Charleston Area Medical Center U54GM104942: West Virginia Clinical and Translational Science Institute (WVCTSI) • Children’s Hospital Colorado — UL1TR002535: Colorado Clinical and Translational Sciences Institute • Columbia University Irving Medical Center — UL1TR001873: Irving Institute for Clinical and Translational Research • Duke University — UL1TR002553: Duke Clinical and Translational Science Institute • George Washington Children’s Research Institute — UL1TR001876: Clinical and Translational Science Institute at Children’s National (CTSA-CN) • George Washington University — UL1TR001876: Clinical and Translational Science Institute at Children’s National (CTSA-CN) • Indiana University School of Medicine — UL1TR002529: Indiana Clinical and Translational Science Institute • Johns Hopkins University — UL1TR003098: Johns Hopkins Institute for Clinical and Translational Research • Loyola Medicine — Loyola University Medical Center • Loyola University Medical Center — UL1TR002389: The Institute for Translational Medicine (ITM) • Maine Medical Center — U54GM115516: Northern New England Clinical & Translational Research (NNE-CTR) Network • Massachusetts General Brigham — UL1TR002541: Harvard Catalyst • Mayo Clinic Rochester UL1TR002377: Mayo Clinic Center for Clinical and Translational Science (CCaTS) • Medical University of South Carolina — UL1TR001450: South Carolina Clinical & Translational Research Institute (SCTR) • Montefiore Medical Center — UL1TR002556: Institute for Clinical and Translational Research at Einstein and Montefiore • Nemours — U54GM104941: Delaware CTR ACCEL Program • NorthShore University HealthSystem — UL1TR002389: The Institute for Translational Medicine (ITM) • Northwestern University at Chicago — UL1TR001422: Northwestern University Clinical and Translational Science Institute (NUCATS) • OCHIN — INV-018455: Bill and Melinda Gates Foundation grant to Sage Bionetworks • Oregon Health & Science University — UL1TR002369: Oregon Clinical and Translational Research Institute • Penn State Health Milton S. Hershey Medical Center — UL1TR002014: Penn State Clinical and Translational Science Institute • Rush University Medical Center — UL1TR002389: The Institute for Translational Medicine (ITM) • Rutgers, The State University of New Jersey — UL1TR003017: New Jersey Alliance for Clinical and Translational Science • Stony Brook

University — U24TR002306 • The Ohio State University — UL1TR002733: Center for Clinical and Translational Science • The State University of New York at Buffalo — UL1TR001412: Clinical and Translational Science Institute • The University of Chicago — UL1TR002389: The Institute for Translational Medicine (ITM) • The University of Iowa — UL1TR002537: Institute for Clinical and Translational Science • The University of Miami Leonard M. Miller School of Medicine — UL1TR002736: University of Miami Clinical and Translational Science Institute • The University of Michigan at Ann Arbor — UL1TR002240: Michigan Institute for Clinical and Health Research • The University of Texas Health Science Center at Houston — UL1TR003167: Center for Clinical and Translational Sciences (CCTS) • The University of Texas Medical Branch at Galveston — UL1TR001439: The Institute for Translational Sciences • The University of Utah — UL1TR002538: Uhealth Center for Clinical and Translational Science • Tufts Medical Center — UL1TR002544: Tufts Clinical and Translational Science Institute • Tulane University — UL1TR003096: Center for Clinical and Translational Science • University Medical Center New Orleans — U54GM104940: Louisiana Clinical and Translational Science (LA CaTS) Center • University of Alabama at Birmingham — UL1TR003096: Center for Clinical and Translational Science • University of Arkansas for Medical Sciences — UL1TR003107: UAMS Translational Research Institute • University of Cincinnati — UL1TR001425: Center for Clinical and Translational Science and Training • University of Colorado Denver, Anschutz Medical Campus — UL1TR002535: Colorado Clinical and Translational Sciences Institute • University of Illinois at Chicago — UL1TR002003: UIC Center for Clinical and Translational Science • University of Kansas Medical Center — UL1TR002366: Frontiers: University of Kansas Clinical and Translational Science Institute • University of Kentucky — UL1TR001998: UK Center for Clinical and Translational Science • University of Massachusetts Medical School Worcester — UL1TR001453: The UMass Center for Clinical and Translational Science (UMCCTS) • University of Minnesota — UL1TR002494: Clinical and Translational Science Institute • University of Mississippi Medical Center — U54GM115428: Mississippi Center for Clinical and Translational Research (CCTR) • University of Nebraska Medical Center — U54GM115458: Great Plains IDeA-Clinical & Translational Research • University of North Carolina at Chapel Hill — UL1TR002489: North Carolina Translational and Clinical Science Institute • University of Oklahoma Health Sciences Center — U54GM104938: Oklahoma Clinical and Translational Science Institute (OCTSI) • University of Rochester — UL1TR002001: UR Clinical & Translational Science Institute • University of Southern California — UL1TR001855: The Southern California Clinical and Translational Science Institute (SC CTSI) • University of Vermont — U54GM115516: Northern New England Clinical & Translational Research (NNE-CTR) Network • University of Virginia — UL1TR003015: iTHRIV Integrated Translational health Research Institute of Virginia • University of Washington — UL1TR002319: Institute of Translational Health Sciences • University of Wisconsin-Madison — UL1TR002373: UW Institute for Clinical and Translational Research • Vanderbilt University Medical Center — UL1TR002243: Vanderbilt Institute for Clinical and Translational Research • Virginia Commonwealth University — UL1TR002649: C. Kenneth and Dianne Wright Center for Clinical and Translational Research • Wake Forest University Health Sciences — UL1TR001420: Wake Forest Clinical and Translational Science Institute • Washington University in St. Louis — UL1TR002345: Institute of Clinical and Translational Sciences • Weill Medical College of Cornell University — UL1TR002384: Weill Cornell Medicine Clinical and Translational Science Center • West Virginia University — U54GM104942: West Virginia Clinical and Translational Science Institute (WVCTSI)

Submitted: Icahn School of Medicine at Mount Sinai — UL1TR001433: ConduITS Institute for Translational Sciences • The University of Texas Health Science Center at Tyler — UL1TR003167: Center for Clinical and Translational Sciences (CCTS) • University of California, Davis — UL1TR001860: UCDavis Health Clinical and Translational Science Center • University of California, Irvine — UL1TR001414: The UC Irvine Institute for Clinical and Translational Science (ICTS) • University of California, Los Angeles — UL1TR001881: UCLA Clinical Translational Science Institute • University of California, San Diego — UL1TR001442: Altman Clinical and Translational Research Institute • University of California, San Francisco — UL1TR001872: UCSF Clinical and Translational Science Institute

Pending: Arkansas Children’s Hospital — UL1TR003107: UAMS Translational Research Institute • Baylor College of Medicine — None (Voluntary) • Children’s Hospital of Philadelphia UL1TR001878: Institute for Translational Medicine and Therapeutics • Cincinnati Children’s Hospital Medical Center — UL1TR001425: Center for Clinical and Translational Science and Training • Emory University — UL1TR002378: Georgia Clinical and Translational Science Alliance • HonorHealth — None (Voluntary) • Loyola University Chicago — UL1TR002389: The Institute for Translational Medicine (ITM) • Medical College of Wisconsin — UL1TR001436: Clinical and Translational Science Institute of Southeast Wisconsin • MedStar Health Research Institute — UL1TR001409: The Georgetown-Howard Universities Center for Clinical and Translational Science (GHUCCTS) • MetroHealth — None (Voluntary) • Montana State University — U54GM115371: American Indian/Alaska Native CTR • NYU Langone Medical Center — UL1TR001445: Langone Health’s Clinical and Translational Science Institute • Ochsner Medical Center — U54GM104940: Louisiana Clinical and Translational Science (LA CaTS) Center • Regenstrief Institute — UL1TR002529: Indiana Clinical and Translational Science Institute • Sanford Research — None (Voluntary) • Stanford University — UL1TR003142: Spectrum: The Stanford Center for Clinical and Translational Research and Education • The Rockefeller University — UL1TR001866: Center for Clinical and Translational Science • The Scripps Research Institute — UL1TR002550: Scripps Research Translational Institute • University of Florida — UL1TR001427: UF Clinical and Translational Science Institute •University of New Mexico Health Sciences Center — UL1TR001449: University of New Mexico Clinical and Translational Science Center • University of Texas Health Science Center at San Antonio — UL1TR002645: Institute for Integration of Medicine and Science • Yale New Haven Hospital — UL1TR001863: Yale Center for Clinical Investigation

## Supplemental Figures

**Figure 1.**
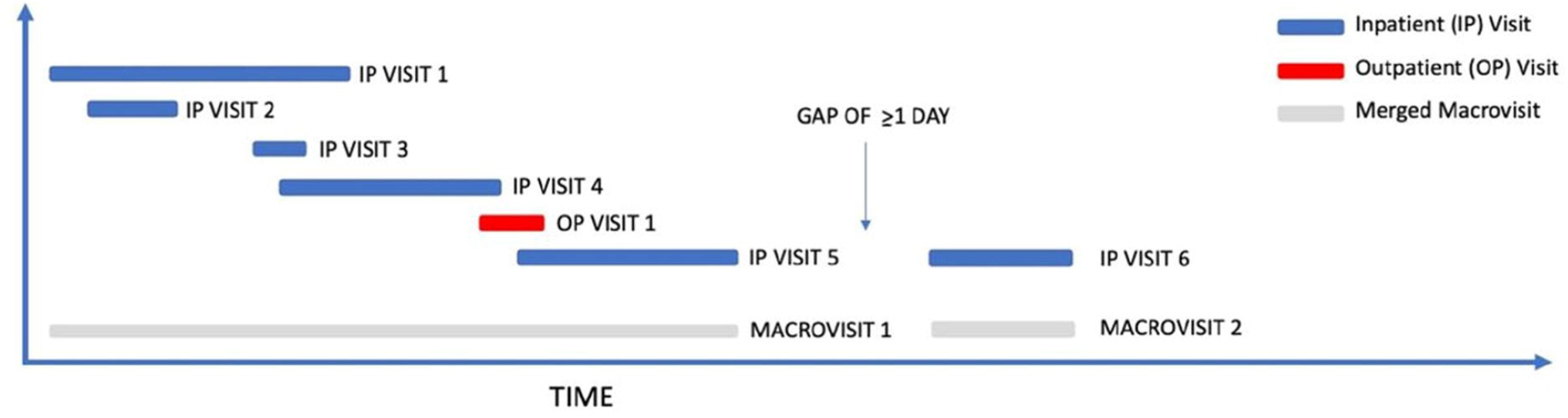
Comparison of LOS distribution between inpatient visits (green bars), macrovisits (red bars), and high-confidence hospitalizations (blue bars) across all sites.

**Figure 2.**
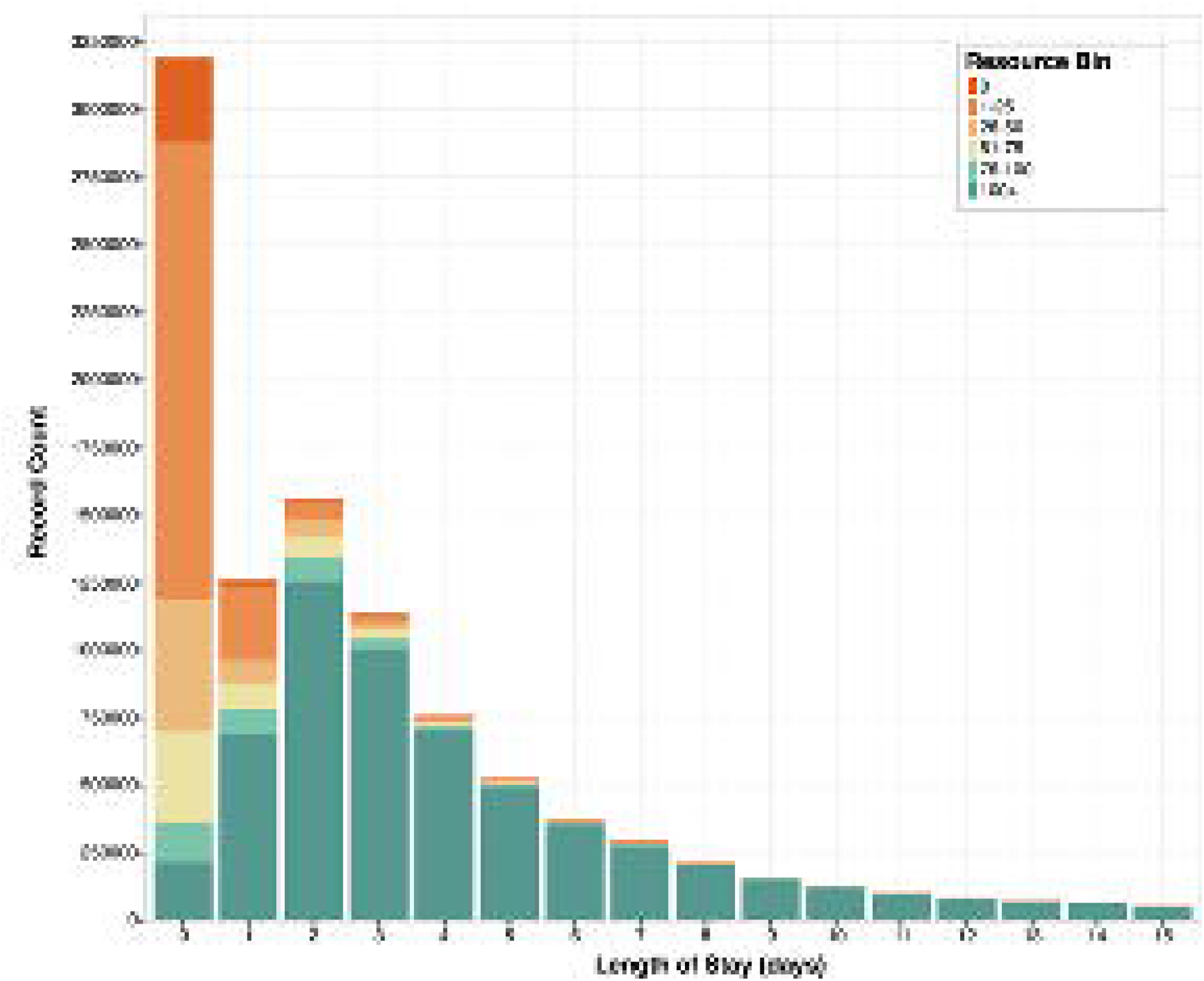
Comparison of cumulative distribution functions showing reduction in LOS variation between macrovists and high-confidence hospitalizations following the macrovisit and high-confidence hospitalization algorithms (sites with largest change noted by color).

**Figure 3.**
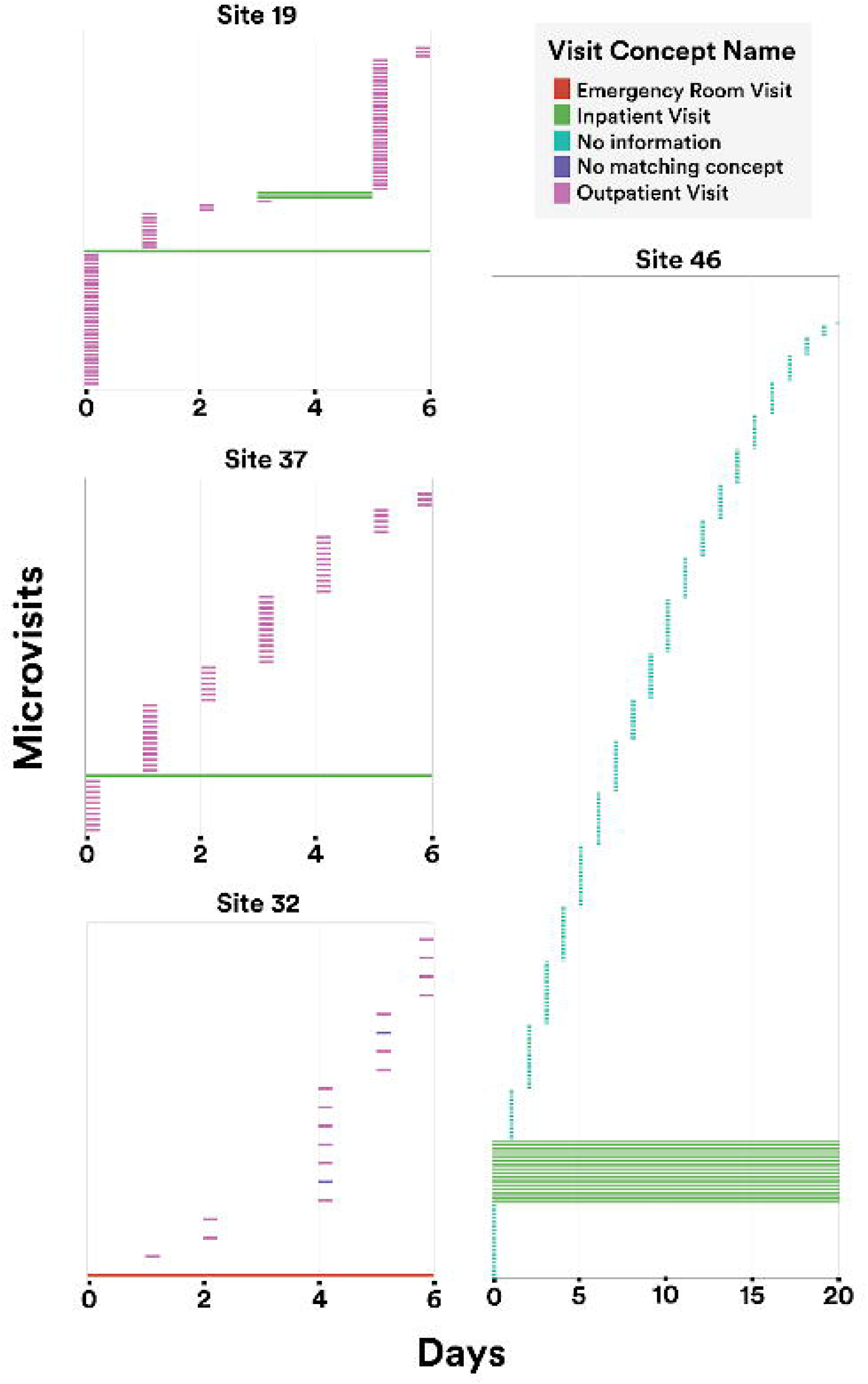
Microvisit heterogeneity within macrovisits across all sites.

**Figure 4.**
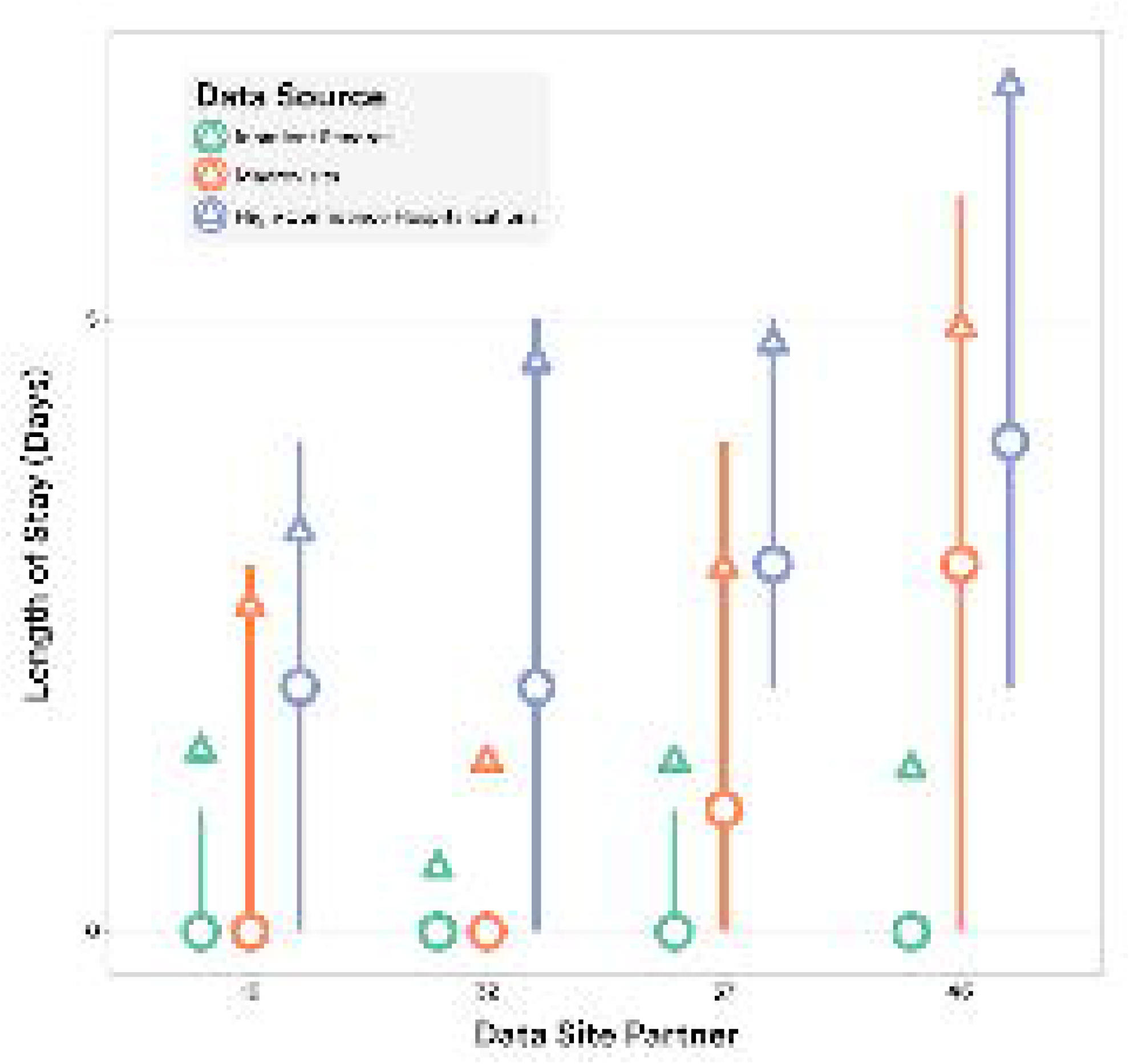
LOS distributions for inpatient visits (blue), macrovisits (orange), and high-confidence hospitalizations (purple) across all sites (median=circle, mean=triangle, IQR=line).

**Figure 5.**
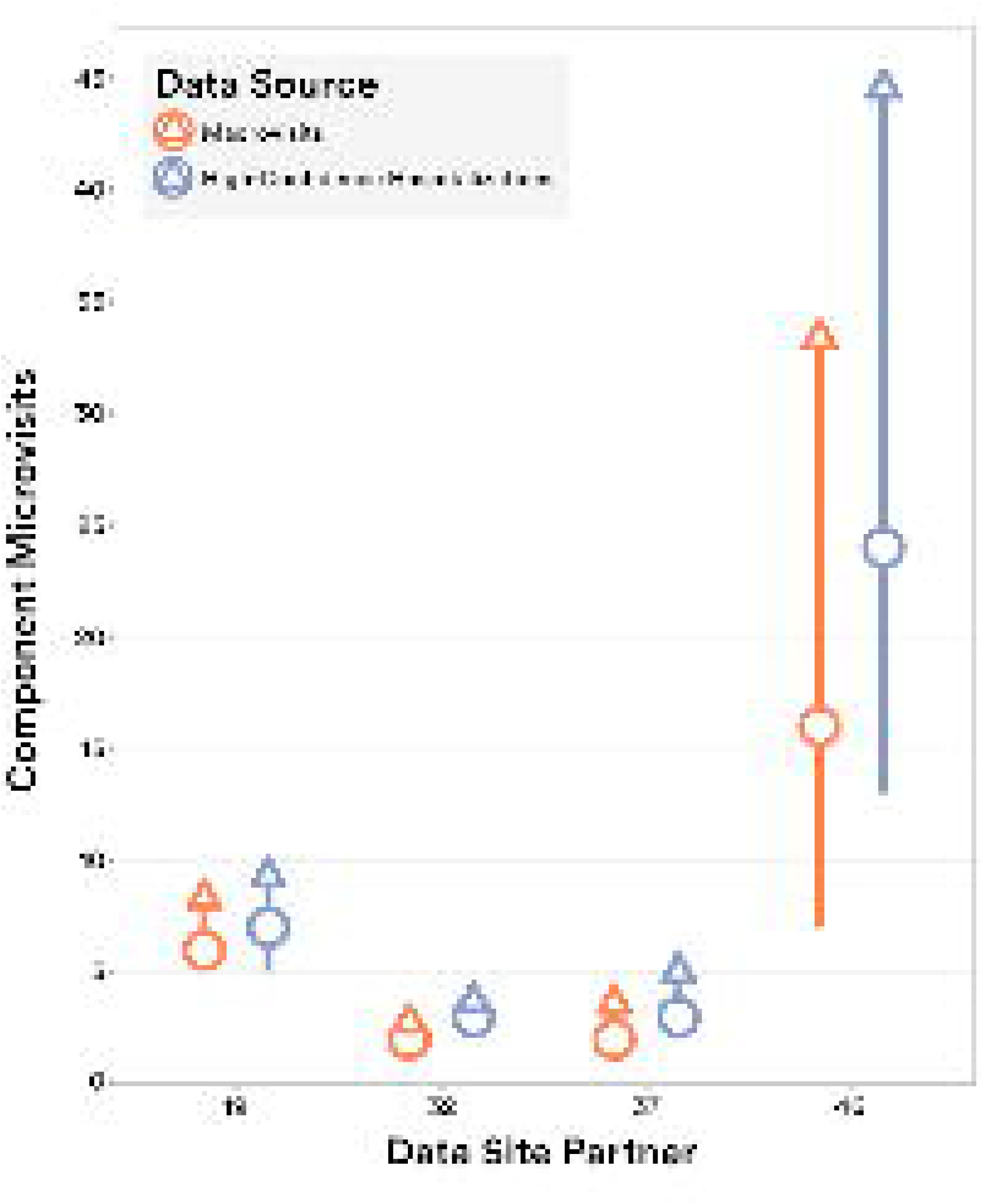
Distribution of the number of measurements within inpatient visits, macrovisits, and high-confidence hospitalizations across all sites (median=circle, mean=triangle, IQR=line).

**Figure 6.**
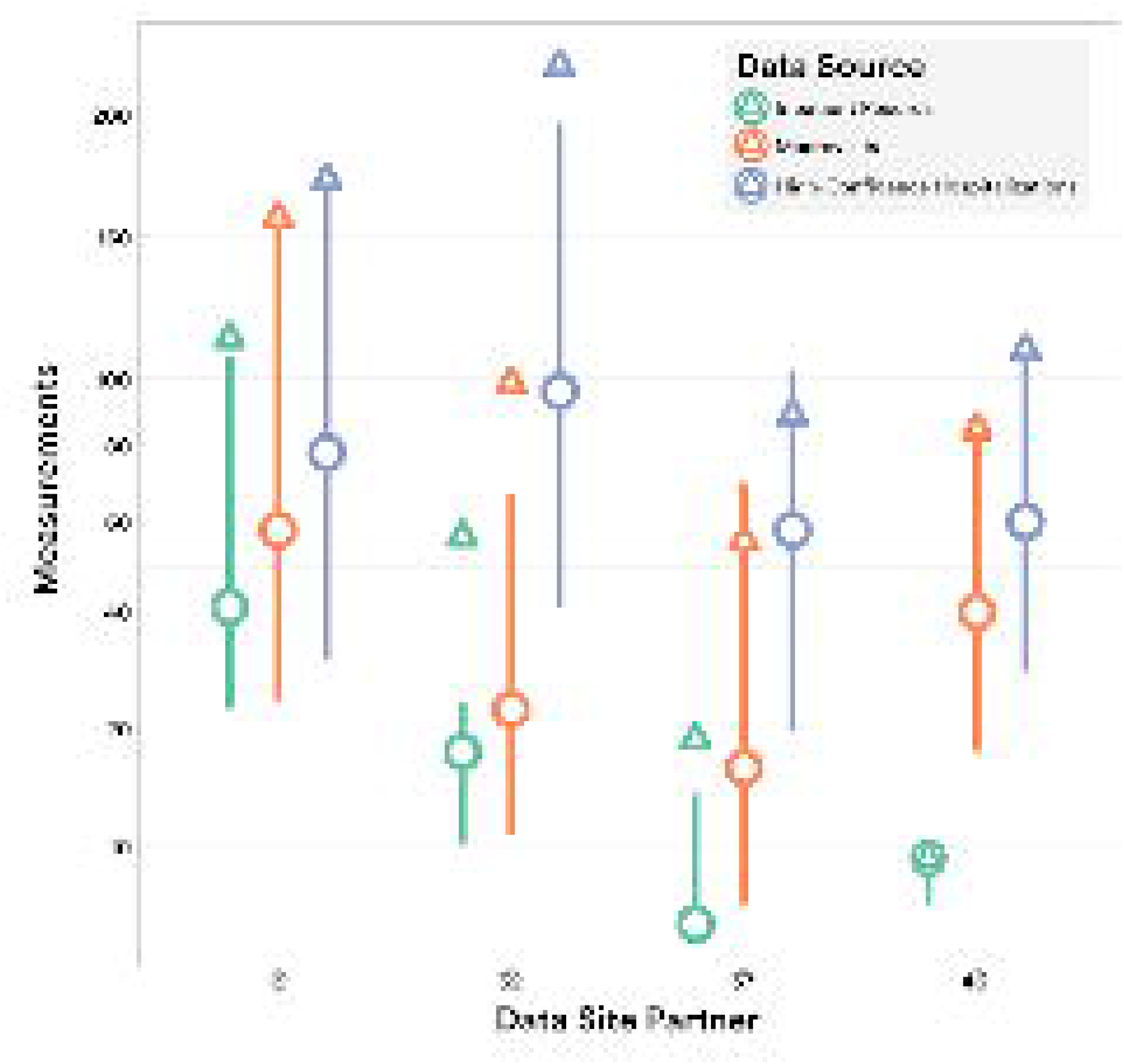
Distribution of the number of component microvisits within macrovisits and high-confidence hospitalizations across all sites (median=circle, mean=triangle, IQR=line).

## Notes

### Competing Interest Statement

The authors have declared no competing interest.

### Funding Statement

This study is part of the NIH Researching COVID to Enhance Recovery (RECOVER)
Initiative, which seeks to understand, treat, and prevent the post-acute sequelae of
SARS-CoV-2 infection (PASC). For more information on RECOVER, visit https://recovercovid.org/. This research was funded by the National Institutes of Health (NIH) Agreement OTA OT2HL161847 as part of the Researching COVID to Enhance Recovery (RECOVER) research program.

### Author Declarations

Ethics committee/IRB of The Johns Hopkins University gave ethical approval for this work

### Summary of Updates

Additional ECMO analysis performed and results added. Several figures were re-formatted to ease reading. The max and min of distributions were move to 1st and 99th percentiles. Minor text edits were performed.

